# Early Prediction of Sepsis in the ICU using Machine Learning: A Systematic Review

**DOI:** 10.1101/2020.08.31.20185207

**Authors:** Michael Moor, Bastian Rieck, Max Horn, Catherine R. Jutzeler, Karsten Borgwardt

## Abstract

**Background:** Sepsis is among the leading causes of death in intensive care units (ICU) world-wide and its recognition, particularly in the early stages of the disease, remains a medical challenge. The advent of an affluence of available digital health data has created a setting in which machine learning can be used for digital biomarker discovery, with the ultimate goal to advance the early recognition of sepsis.

**Objective:** To systematically review and evaluate studies employing machine learning for the prediction of sepsis in the ICU.

**Data sources:** Using Embase, Google Scholar, PubMed/Medline, Scopus, and Web of Science, we systematically searched the existing literature for machine learning-driven sepsis onset prediction for patients in the ICU.

**Study eligibility criteria:** All peer-reviewed articles using machine learning for the prediction of sepsis onset in adult ICU patients were included. Studies focusing on patient populations outside the ICU were excluded.

**Study appraisal and synthesis methods:** A systematic review was performed according to the PRISMA guidelines. Moreover, a quality assessment of all eligible studies was performed.

**Results:** Out of 974 identified articles, 22 and 21 met the criteria to be included in the systematic review and quality assessment, respectively. A multitude of machine learning algorithms were applied to refine the early prediction of sepsis. The quality of the studies ranged from “poor” (satisfying ≤ 40% of the quality criteria) to “very good” (satisfying ≥ 90% of the quality criteria). The majority of the studies (*n* = 19, 86.4%) employed an offline training scenario combined with a horizon evaluation, while two studies implemented an online scenario (*n* = 2, 9.1%). The massive inter-study heterogeneity in terms of model development, sepsis definition, prediction time windows, and outcomes precluded a meta-analysis. Last, only 2 studies provided publicly-accessible source code and data sources fostering reproducibility.

**Limitations:** Articles were only eligible for inclusion when employing machine learning algorithms for the prediction of sepsis onset in the ICU. This restriction led to the exclusion of studies focusing on the prediction of septic shock, sepsis-related mortality, and patient populations outside the ICU.

**Conclusions and key findings:** A growing number of studies employs machine learning to optimise the early prediction of sepsis through digital biomarker discovery. This review, however, highlights several shortcomings of the current approaches, including low comparability and reproducibility. Finally, we gather recommendations how these challenges can be addressed before deploying these models in prospective analyses.

**Systematic review registration number:** CRD42020200133

## 1 Introduction

Sepsis is a life-threatening organ dysfunction triggered by dysregulated host response to infection [Singer et al., 2016] and constitutes a major global health concern [Rudd et al., 2020]. Despite promising medical advances over the last decades, sepsis remains among the most common causes of in-hospital deaths. It is associated with an alarmingly high mortality and morbidity, and massively burdens the health care systems world-wide [Dellinger et al., 2013, Hotchkiss et al. 2016a, Kaukonen et al., 2014, Rudd et al., 2020]. In parts, this can be attributed to challenges related to early recognition of sepsis and initiation of timely and appropriate treatment [Ferrer et al. 2014]. A growing number of studies suggests that the mortality increases with every hour the antimicrobial intervention is delayed—further underscoring the importance of timely recognition and initiation of treatment [Ferrer et al., 2014, Pruinelli et al., 2018, Weiss et al., 2014]. A major challenge to early recognition is to distinguish sepsis from disease states (e.g. inflammation) that are hallmarked by similar clinical signs (e.g. change in vitals), symptoms (e.g. fever), and molecular manifestations (e.g. dysregulated host response) [Al Jalbout et al., 2019, Lever and Mackenzie 2007]. Owing to the systemic nature of sepsis, biological and molecular correlates—also known as biomarkers—have been proposed to refine the diagnosis and detection of sepsis [Hotchkiss et al., 2016b]. However, despite considerable efforts to identify suitable biomarkers, there is yet no single biomarker or set thereof that is universally accepted for sepsis diagnosis and treatment, mainly due to the lack of sensitivity and specificity [Faix, 2013, Parlato et al., 2018].

In addition to the conventional approaches, *data-driven* biomarker discovery has gained momentum over the last decades and holds the promise to overcome existing hurdles. The goal of this approach is to mine and exploit health data with quantitative computational approaches, such as machine learning. An ever-increasing amount of data, including laboratory, vital, genetic, molecular, as well as clinical data and health history, is available in digital form and at high resolution for individuals at risk and for patients suffering from sepsis [Johnson et al., 2016b]. This versatility of the data allows to search for digital biomarkers in a holistic fashion as opposed to a reductionist approach (e.g. solely focusing on hematological markers). Machine learning models can naturally handle the wealth and complexity of digital patient data by learning predictive patterns in the data, which in turn can be used to make accurate predictions about which patient is developing sepsis [Fleuren et al., 2020, Thorsen-Meyer et al., 2020]. Over the last decades, multiple studies have successfully employed a variety of computational models to tackle the challenge of predicting sepsis at the earliest time point possible [Barton et al., 2019, Kaji et al., 2019, McCoy and Das, 2017]. For instance, Futoma and colleagues proposed to combine multi-task Gaussian processes imputation together with a recurrent neural network in one end-to-end trainable framework (MGP-RNN). They were able to predict sepsis 17h prior to the first administration of antibiotics and 36h before a definition for sepsis was met [Futoma et al., 2017b]. This strategy was motivated by Li and Marlin [2016], who first proposed the so-called Gaussian process adapter that combines single-task Gaussian processes imputation with neural networks in an end-to-end learning setting. A more recent study further improved predictive performance by combining the Gaussian process adapter framework with temporal convolutional networks (MGP-TCN) as well as leveraging a dynamic time warping approach for the early prediction of sepsis [Moor et al., 2019].

Considering the rapid pace at which the research in this field is moving forward, it is important to summarise and critically assess the state of the art. Thus, the aim of this review. Thus, the aim of this review was to provide a comprehensive overview of the current state of machine learning models that have been employed for the search of digital biomarkers to aid the early prediction of sepsis in the intensive care unit (ICU). To this end, we systematically reviewed the literature and performed a quality assessment of all eligible studies. Based on our findings, we also provide some recommendations for forthcoming studies that plan to use machine learning models for the early prediction of sepsis.

## 2 Methods

The study protocol was registered with and approved by the international prospective register of systematic reviews (PROSPERO) before the start of the study (registration number: CRD42020200133). We followed the Preferred Reporting Items for Systematic reviews and Meta-Analysis (PRISMA) statement [Moher et al., 2015]

### 2.1 Search strategy and selection criteria

Five bibliographic databases were systematically searched, i.e. EMBASE, Google Scholar, PubMed/Medline, Scopus, and Web of Science, using the time range from their respective inception dates to July 20th, 2020. Google Scholar was searched using the tool “Publish or Perish” (version 7.23.2852.7498) [Harzing, 2007]. Our search was not restricted by language. The search term string was constructed as (“sepsis prediction” OR “sepsis detection”) AND (“machine learning” OR “artificial intelligence”) to include publications focusing on (early) onset prediction of sepsis with different machine learning methods. The full search strategy is provided in Supplementary Table 1.

### 2.2 Selection of studies

Two investigators (MM and CRJ) independently screened the titles, abstracts, and full-texts retrieved from Google Scholar in order to determine the eligibility of the studies. Google Scholar was selected by virtue of its promise of an inclusive query that also captures conference proceedings, which are highly relevant to the field of machine learning but not necessarily indexed by other databases. In a second step, two investigators (MM and MH) queried EMBASE, PubMed, Scopus, and Web of Science for additional studies. Eligibility criteria were also applied to the full-text articles during the final selection. In case multiple articles reported on a single study, the article that provided the most data and details was selected for further synthesis. We quantified the inter-rater agreement for study selection using Cohen’s kappa (*κ*) coefficient [Viera et al., 2005]. All disagreements were discussed and resolved at a consensus meeting.

### 2.3 Inclusion and exclusion criteria

All full-text, peer-reviewed articles^1^ using machine learning for the prediction of sepsis onset in the ICU were included. Although the 2016 consensus statement abandoned the term “severe sepsis” [Singer et al., 2016], studies published prior to the revised consensus statement targeting severe sepsis were also included in our review. Furthermore, to be included, studies must have provided sufficient information on the machine learning algorithms used for the analysis, definition of sepsis (e.g. Sepsis-3), and sepsis onset definition (e.g. time of suspicion of infection). We excluded duplicates, non-peer reviewed articles (e.g. preprints), reviews, meta-analyses, abstracts, editorials, commentaries, perspectives, patents, letters with insufficient data, studies on non-human species and children/neonates, or out-of-scope studies (e.g. different target condition). Lastly, studies focusing on the prediction of septic shock were also excluded as the septic shock was beyond the scope of this review. The extraction was performed by four investigators (MM, BR, MH, and CRJ).

### 2.4 Data extraction and synthesis

The following information was extracted from all studies: (1) publication characteristics (first author’s last name, publication time), (2) study design (retrospective, prospective data collection and analysis), (3) cohort selection (sex, age, prevalence of sepsis), (4) model selection (machine learning algorithm, platforms, software, packages, and parameters), (5) specifics on the data analysed (type of data, number of variables), (6) statistics for model performance (methods to evaluate the model, mean, measure of variance, handling of missing data), and (7) methods to avoid overfitting as well as any additional external validation strategies. If available, we also reviewed supplementary materials of each study. A full list of extracted variables is provided in Supplementary Table 2.

### 2.5 Settings of prediction task

Owing to its time-sensitivity, setting up the early sepsis prediction task in a clinically-meaningful manner is a non-trivial issue. We extracted details on the prediction task as well as the alignment of cases and controls. Given the lack of standardised reporting, the implementation strategies and their reporting vary drastically between studies. Thus, subsequent to gathering all the information, we attempted to create new categories for the sepsis prediction task as well as the case-control alignment. The goal of this new terminology and categories is to increase the comparability between studies.

### 2.6 Assessment of quality of reviewed machine learning studies

Based on 14 criteria relevant to the objectives of the review (adapted from Qiao [2019]), the quality of the eligible machine learning studies was assessed. The quality assessment comprised five categories: (1) unmet needs (limits in current machine learning or non-machine learning applications), (2) reproducibility (information on the sepsis prevalence, data and code availability, explanation of sepsis label, feature engineering methods, software/hardware specifications, and hyperparameters), (3) robustness (sample size suited for machine learning applications, valid methods to overcome overfitting, stability of results), (4) generalisability (external data validation), and (5) clinical significance (interpretation of predictors and suggested clinical use; see Supplementary Table 3). A quality assessment table was provided by listing “yes” or “no” of corresponding items in each category. MM, BR, MH, and CRJ independently performed the quality assessment. In case of disagreements, ratings were discussed and subsequently, final scores for each publication were determined.

## 3 Results

### 3.1 Study selection

The results of the literature search, including the numbers of studies screened, assessments for eligibility, and articles reviewed (with reasons for exclusions at each stage), are presented in Figure 1. Out of 974 studies, 22 studies met the inclusion criteria [Abromavičius et al., 2020, Barton et al., 2019, Bloch et al. 2019, Calvert et al., 2016, Desautels et al., 2016, Futoma et al., 2017b, Kaji et al., 2019, Kam and Kim, 2017, Lauritsen et al., 2020, Lukaszewski et al., 2008, Mao et al., 2018, McCoy and Das, 2017, Moor et al., 2019, Nemati et al. 2018, Reyna et al., 2019, Schamoni et al., 2019, Scherpf et al., 2019, Shashikumar et al., 2017a,b, Sheetrit et al., 2019, Van Wyk et al., 2019, van Wyk et al., 2019]. The majority of excluded studies (*n* = 952) did not meet one or multiple inclusion criteria, such as studying a non-human (e.g. bovine) or a non-adult population (e.g. paediatric or neonatal), focusing on a research topic beyond the current review (e.g. sepsis phenotype identification or mortality prediction), or following a different study design (e.g. case reports, reviews, not-peer reviewed). Detailed information on all included studies are provided in Table 1. The inter-rater agreement was excellent (*κ* = 0.88).

**Figure 1:**
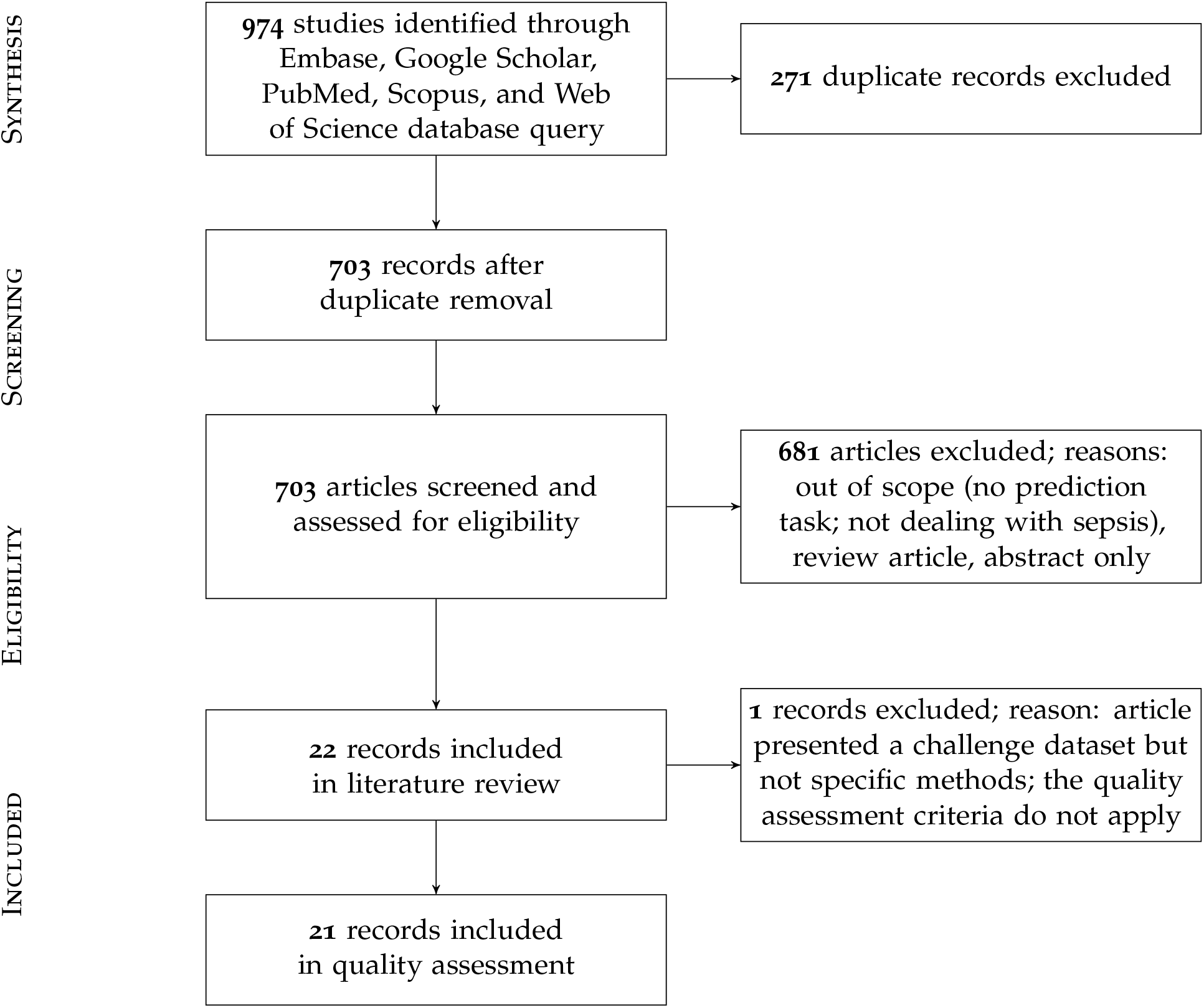
PRISMA flowchart of the search strategy. A total of 22 studies were eligible for the literature review and 21 for the quality assessment.

**Table 1:**
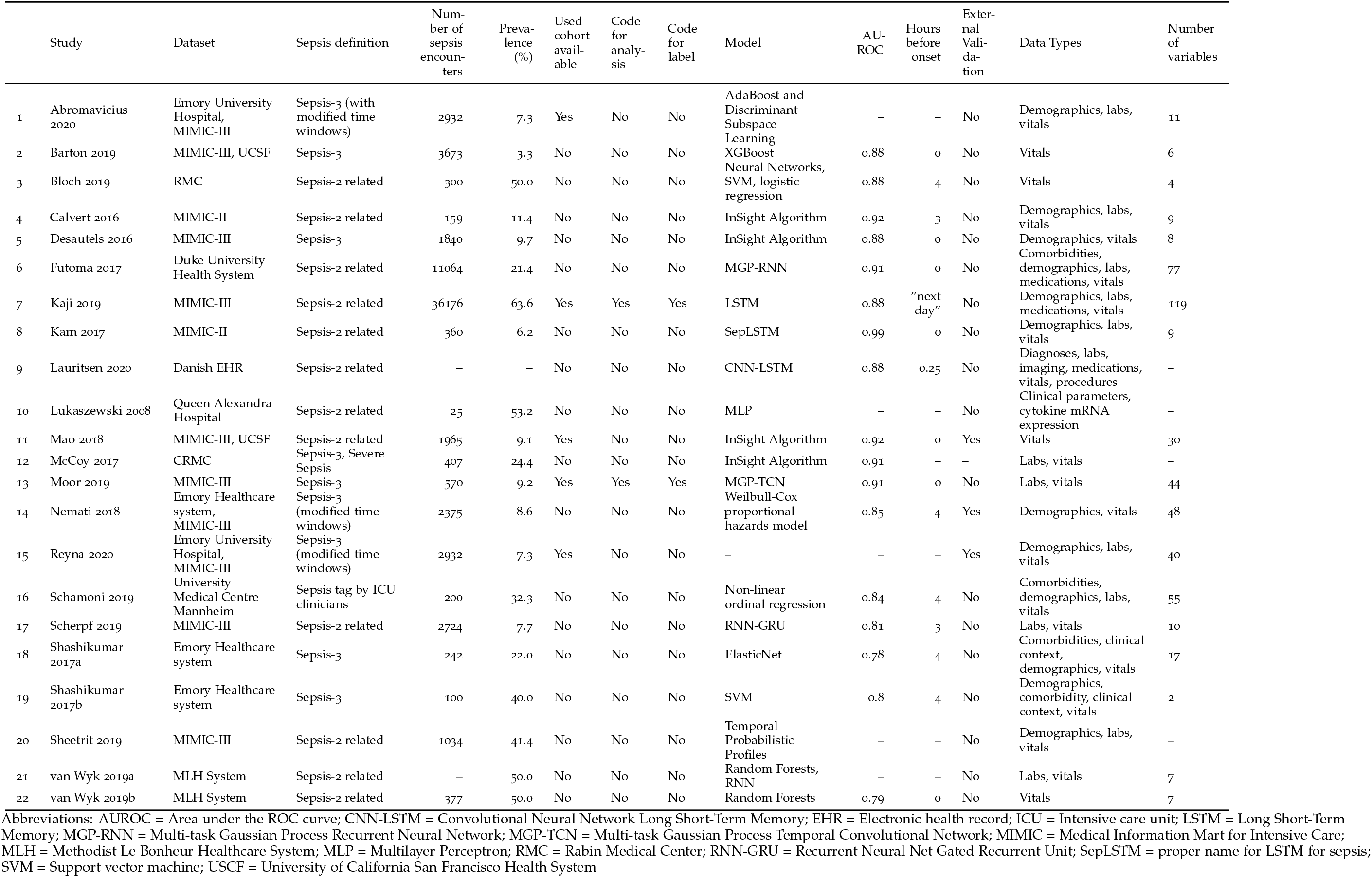
Overview of included studies. Only if area under the Receiver Operating Characteristic Curve (AUROC) was reported in an early prediction setup, the performance and the corresponding prediction window is reported (in hours before onset). As these windows were highly heterogeneous, to achieve more comparability, we report the minimal hour before onset that was reported. Notably, due to heterogeneous sepsis definition implementations and experimental setups, these metrics likely have low comparability between studies, which is why we deemed a quantitative meta-analysis to be inappropriate.

### 3.2 Study characteristics

Of the 22 included studies, 21 employed solely retrospective analyses, while 1 study used both retrospective and prospective analyses [McCoy and Das, 2017]. Moreover, the most frequent data sources used to develop computational models were MIMIC-II and MIMIC-III (*n* = 12; 54.5%), followed by Emory University Hospital (*n* = 5; 22.7%). In terms of sepsis definition, the majority of the studies employed the Sepsis-2 (*n* = 12; 54.5%) or Sepsis-3 definition (*n* = 9; 40.9%). It is important to note that some studies *modified* the Sepsis-2 or Sepsis-3 definition since all existing definitions have not been intended to specify an exact sepsis onset time (e.g. the employed time window lengths have been varied) [Abromavičius et al., 2020, Nemati et al., 2018]. In one study [Schamoni et al., 2019], sepsis labels were assigned by trained ICU experts. Depending on the definition of sepsis used, and whether subsampling of controls was used to achieve a more balanced class ratio (facilitating the training of machine learning models), the prevalence of patients developing sepsis ranged between 6.2% and 63.6% (Figure 2). One study did not report the prevalence [Lauritsen et al., 2020]. Concerning demographics, 9 studies reported the median or mean age, 12 the prevalence of female patients, and solely 1 the ethnicity of the investigated cohorts (Supplementary Table 4).

**Figure 2:**
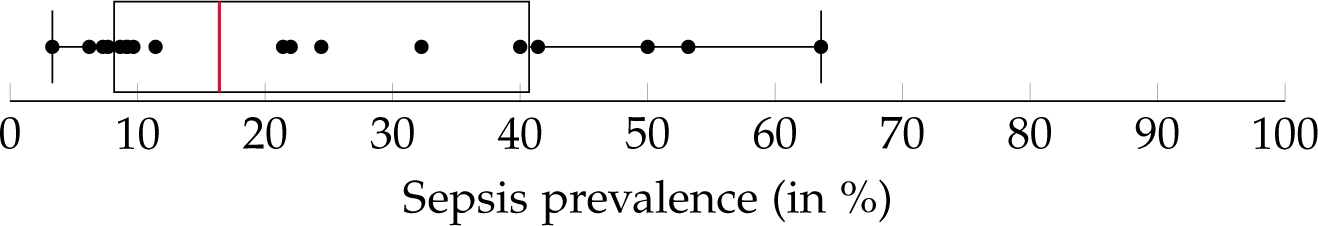
A boxplot of the sepsis prevalence distribution of all studies, with the median prevalence being highlighted in red. Note that some studies have subset controls for balancing the class ratios in order to facilitate the training of the machine learning model. Thus, the prevalence in the study cohort (i.e. the subset) can be different from the prevalence of the original data source (e.g. MIMIC-III).

### 3.3 Overview of machine learning algorithms and data

As shown in Table 1, a wide range of predictive models was employed for the early detection of sepsis, with some models being specifically developed for the respective application. Most prominently, various types of neural networks (*n* = 9; 40.9%) were used. This includes recurrent architectures such as long short-term memory (LSTM) [Hochreiter and Schmidhuber, 1997] or gated recurrent units (GRU) [Cho et al., 2014], convolutional networks [Fukushima et al., 1983], as well as temporal convolutional networks, featuring causal, dilated convolutions [Lea et al. 2017, Oord et al., 2016]. Furthermore, several studies employed boosted tree models (*n* = 4; 18.2%), including XGBoost [Chen and Guestrin, 2016], or random forest [Kam et al., 1995]. As for the data analysed, the most common data type were vitals (*n* = 21; 95.5%), followed by laboratory values (*n* = 13; 59.1%), demographics (*n* = 12; 54.5%), and comorbidities (*n* = 4; 18.2%). The number of variables included in the respective models ranged between 2 [Shashikumar et al. 2017a] and 119 [Kaji et al., 2019]. While reporting the type of variables, four studies failed to Lukaszewski et al. 2020 report the number of variables included in the models [Lauritsen et al. 2008, McCoy and Das, 2017, Sheetrit et al., 2019].

### 3.4 Model validation

Approximately 80% of the studies employed one type of cross-validation (e.g. 5-fold, 10-fold, or leave-one-out cross-validation) to avoid overfitting. Additional validation of the models on out-of-distribution ICU data (i.e. external validation) was only performed in three studies [Mao et al., 2018, Nemati et al., 2018, Reyna et al., 2019]. Specifically, Mao et al. [2018] used a dataset provided by the UCSF Medical Center as well as the MIMIC-III data set to train, validate, and test the *InSight* algorithm. Aiming at developing and validating the *Artificial Intelligence Sepsis Expert (AISE)* algorithm, Nemati et al. [2018] created a development cohort using ICU data of over 30,000 patients admitted to two Emory University hospitals. In a subsequent step, the *AISE* algorithm was externally validated on the publicly-available MIMIC-III dataset (at the time containing data from over 52,000 ICU stays of more than 38,000 unique patients) [Nemati et al., 2018]. Last, the study by Reyna et al. [2019], describes the protocol and results of the *PhysioNet/Computing in Cardiology Challenge 2019*. Briefly, the aim of this challenge was to facilitate the development of automated, open-source algorithms for the early detection of sepsis. The PhysioNet/Computing in Cardiology Challenge provided sequestered real-world datasets to the participating researchers for the training, validation, and testing of their models.

### 3.5 Experimental design choices for sepsis onset prediction

In this review, we identified two main approaches of implementing sepsis prediction tasks on ICU data. The most-frequent setting (*n* = 19; 86.4%) combines “offline” training with a “horizon” evaluation. Briefly, offline training refers to the fact that the models have access to the entire feature window of patient data. For patients sustaining a sepsis, this feature window ranges from hospital admission to sepsis onset, while for the control subjects the endpoint is a matched onset. Alternatively, a prediction window (i.e. a *gap*) between the feature window and the (matched) onset has been employed [Bloch et al., 2019]. As for the “horizon” evaluation, the purpose is to determine how early the fitted model would recognise sepsis. To this end, all input data gathered up to *n* hours before onset is provided to the model for the sepsis prediction at an horizon of *n* hours. For studies employing only a single horizon, i.e. predictions preceding sepsis onset by a fixed number of hours, we denote their task as “offline” evaluation in Table 2, since there are no sequentially repeated predictions over time. This experimental setup, offline training plus horizon evaluation, is visualised in Figure 3. In the second most-frequently used sepsis prediction setting (n = 2; 9.1%), both the training and evaluation occur in an “online” fashion. This means that the model is presented with all the data that has been collected until the time point of prediction. The amount of data depends on the spacing of data collection. In order to incentivise early predictions, these timepoint-wise labels can be shifted into the past: in the case of the PhysioNet Challenge dataset, already timepoint-wise labels 6 h before onset are assigned to the positive (sepsis) class [Reyna et al., 2019]. For an illustration of an online training and evaluation scenario, refer to Figure [4]

**Figure 3:**
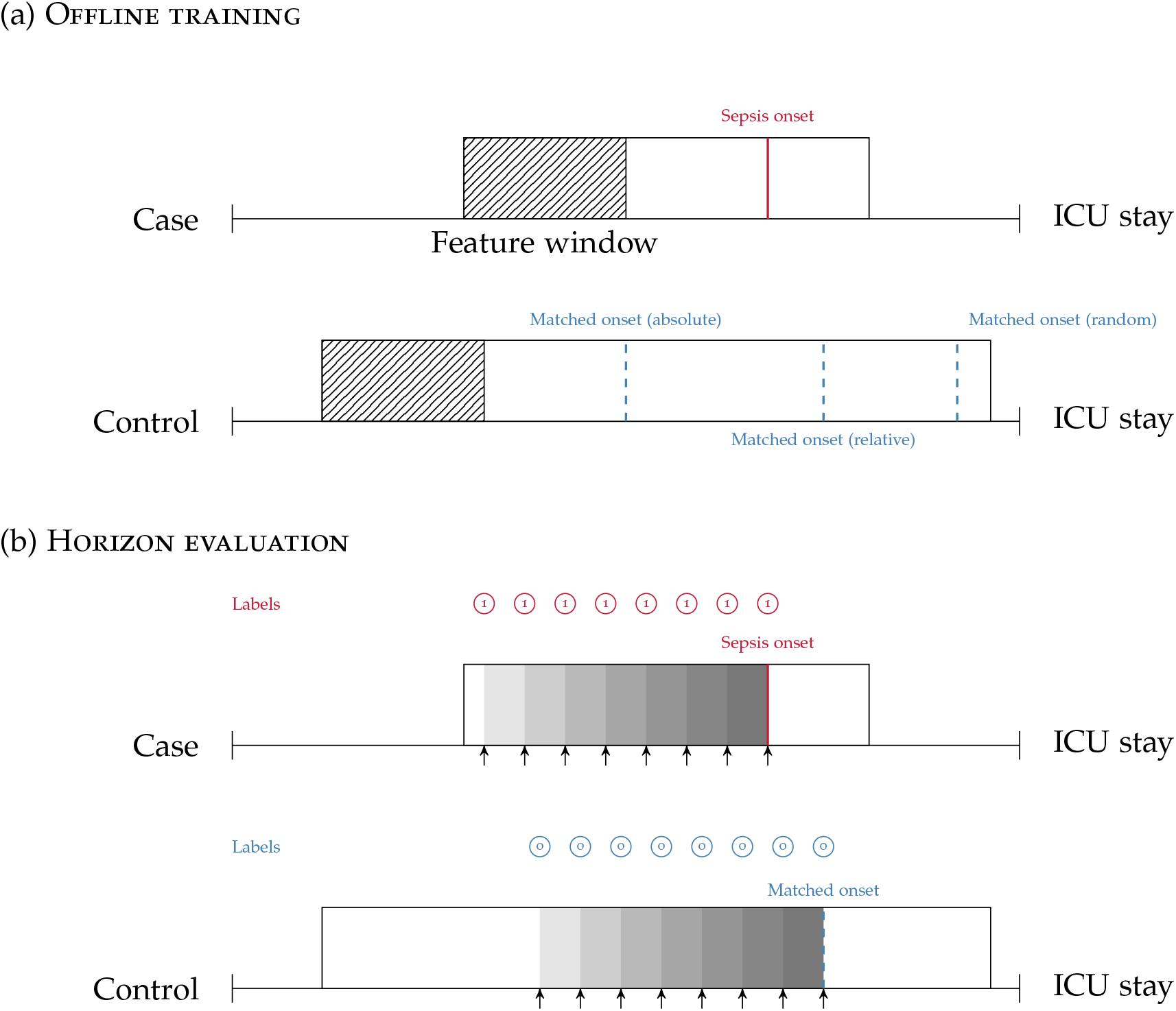
(a): Offline training scenario and case-control matching. Every case has a specific sepsis onset. Given a random control, there are multiple ways of determining a matched onset time: (i) *relative* refers to the relative time since intensive care unit (ICU) admission (here, 75% of the ICU stay); (ii) *absolute* refers to the absolute time since ICU admission; (iii) *random* refers to a pseudo-random time during the ICU stay, often with the requirement that the onset is not too close to ICU discharge. (b): Horizon evaluation scenario. Given a case and control, with a matched relative sepsis onset, the look-back horizon indicates how *early* a specific model is capable of predicting sepsis. As the (matched) sepsis onset is approached, this task typically becomes progressively easier. Notice the difference in the prediction targets (labels) (shown in red for predicting a case, and blue for predicting a control.)

**Table 2:**
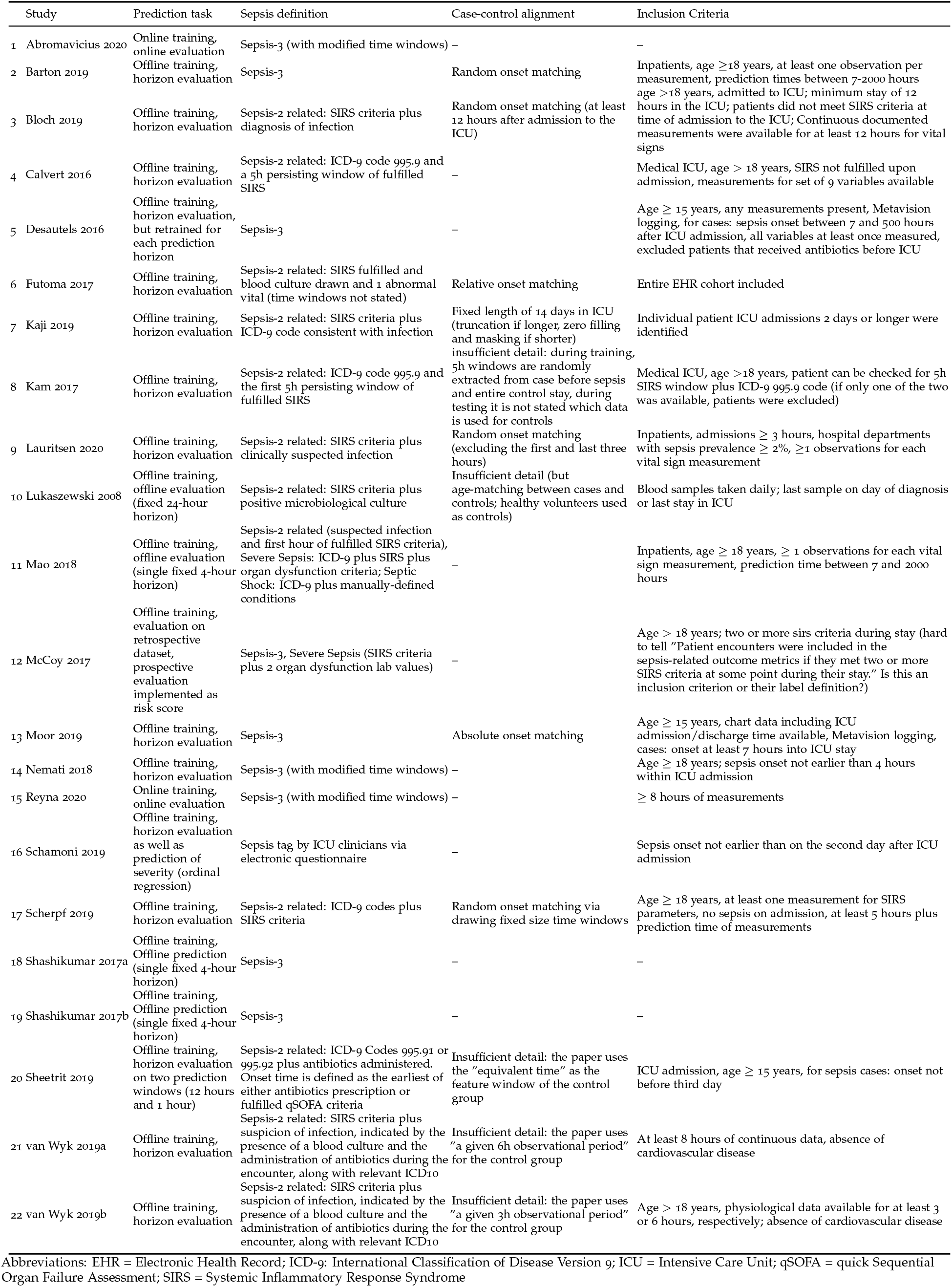
An overview of experimental details: The used sepsis definition, the exact prediction task, and which type of temporal case-control alignment was used (if any).

**Figure 4:**
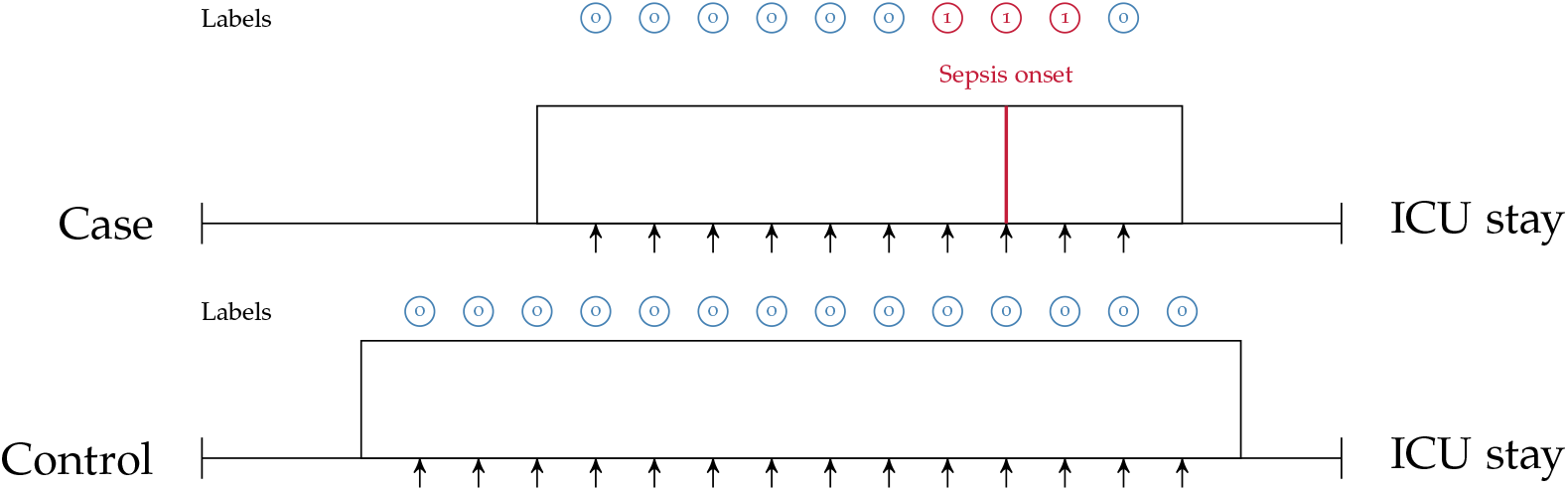
Online training and evaluation scenario. Here, the model predicts at regular intervals during an ICU stay (we show predictions in *1 h* intervals). For sepsis cases, there is no prima facie notion at which point in time positive predictions ought to be considered as true positive (TP) predictions or false positive (FP) predictions (mutatis mutandis, this applies to negative predictions). For illustrative purposes, here we consider positive predictions up until *1 h* before or after sepsis onset (for a case) to be TP.

Selecting the “onset” for controls (i.e. case-control alignment) is a crucial step in the development of models predicting a sepsis onset [Futoma et al., 2017b]. Surprisingly, the majority of the studies (n = 16; 72.7%) did not report any details on how the onset matching was performed. For the six studies (27.3%) providing details, we propose the following classification: four employed *random onset matching*, one *absolute onset matching*, and one *relative onset matching* (Figure 3, top). As the name indicates, during random onset matching, the onset time of a control is set at a random time of the ICU stay. Often, this time has to satisfy certain additional constraints, such as not being too close to the patient’s discharge. The absolute onset matching refers to taking the absolute time since admission until sepsis onset for the case and assigning it as the matched onset time for a control [Moor et al., 2019]. Lastly, the relative onset matching is when the matched onset time is defined as the relative time since ICU admission until sepsis onset for the case [Futoma et al., 2017a].

### 3.6 Quality of included studies

The results of the quality assessment are shown in Table 3. One study [Reyna et al., 2019], showcasing the results of the *PhysioNet/Computing in Cardiology Challenge 2019*, was excluded from the Quality Assessment, which was intended to assess the quality of the implementation and reporting of *specific* prediction models. The quality of the remaining 21 studies ranged from poor (satisfying ≤ 40% of the quality criteria) to very good (satisfying ≥ 90% of the quality criteria). None of the studies fulfilled all 14 criteria. A single criterion was met by 100% of the studies: all studies highlighted the limits in current non-machine-learning approaches in the introduction. Few studies provided the code used for the data cleaning and analysis (*n* = 2; 9.5%), provided data or code for the reproduction of the exact sepsis labels and onset times (*n* = 2; 9.5%), and validated the machine learning models on an external data set (*n* = 3; 14.3%). For the interpretation, power, and validity of machine learning methods, considerable sample sizes are required. With the exception of one study [Lukaszewski et al., 2008], all studies had sample sizes larger than 50 sepsis patients.

**Table 3:**
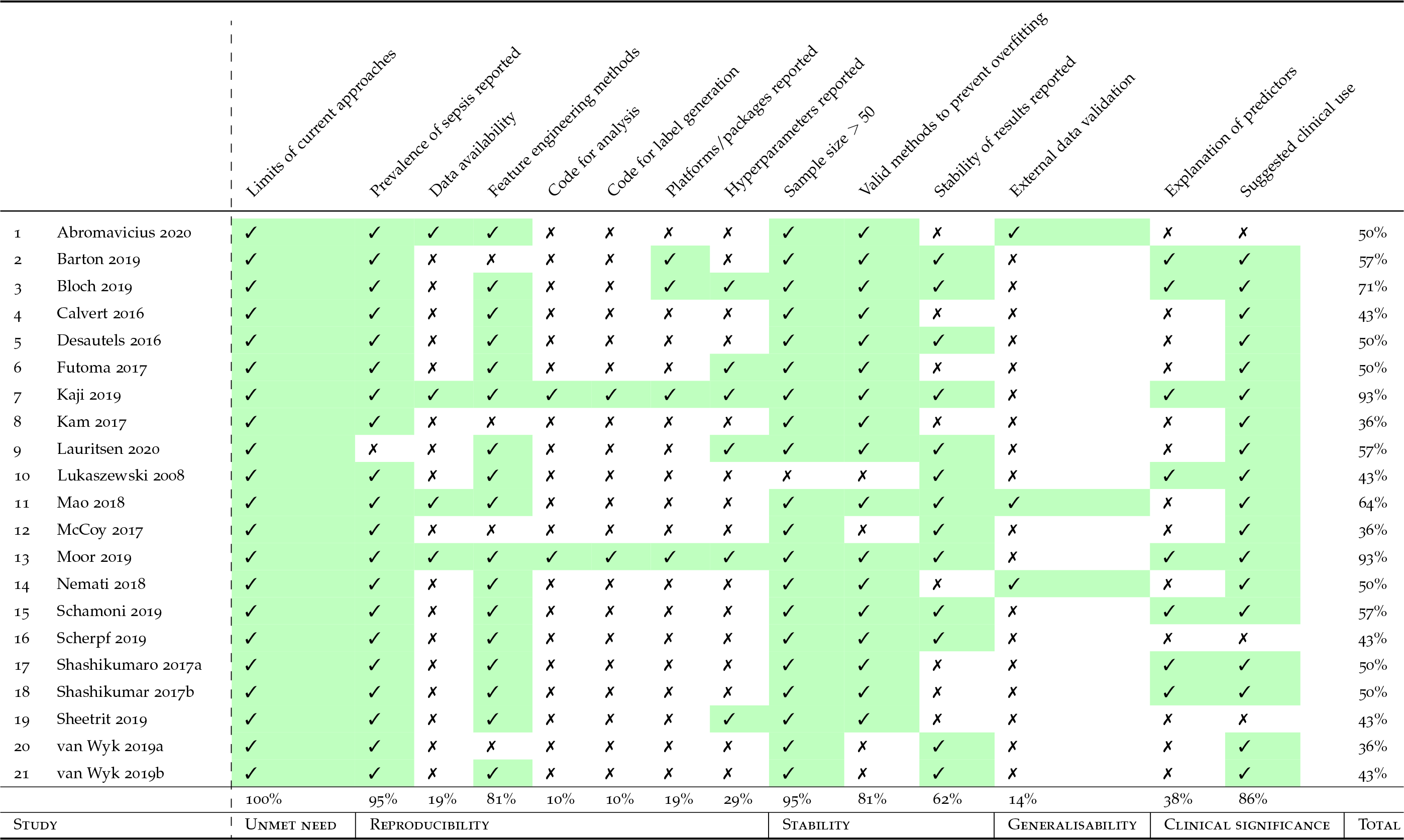
Quality assessment of all studies. We excluded Reyna et al. [2019] from the assessment because it does presents a dataset challenge rather than a single method, making most of the categories not applicable.

## 4 Discussion

In this study, we systematically reviewed the literature for studies employing machine learning algorithms to facilitate early prediction of sepsis. A total of 22 studies were deemed eligible for the review and 21 were included in the quality assessment. The majority of the studies used data from the MIMIC-III database [Johnson et al., 2016b], containing deidentified health data associated with ≈ 60,000 intensive care unit admissions and/or data from Emory University Hospital^2^). With the exception of one, *all* studies used internationally-acknowledged guidelines for sepsis definitions, namely Sepsis-2 [Levy et al., 2003] and Sepsis-3 [Singer et al., 2016]. In terms of the analysis, a wide range of machine learning algorithms were chosen to leverage the patients’ digital health data for the prediction of sepsis. Driven by our findings from the reviewed studies, this section first highlights four major challenges that the literature on machine learning driven sepsis prediction is currently facing: (i) asynchronicity, (ii) comparability, (iii) reproducibility, and (iv) circularity. We then discuss the limitations of this study, provide some recommendations for forthcoming studies, and conclude with an outlook.

### 4.1 Asynchronicity

While initial studies employing machine learning for the prediction of sepsis have demonstrated promising results [Calvert et al., 2016, Desautels et al., 2016, Kam and Kim, 2017], the literature since has been diverging on which are the most pressing open challenges that need to be addressed to further the goal of early sepsis detection. On the one hand, corporations have been propelling the deployment of the first interventional studies [Burdick et al., 2020, Shimabukuro et al., 2017], while on the other hand, recent findings have cast doubt on the validity and meaningfulness of the experimental pipeline that is currently being implemented in most retrospective analyses [Schamoni et al., 2019]. This can be partially attributed to circular prediction settings (for more details, please refer to Section 4.4). Ultimately, only the demonstration of favourable outcomes in large prospective randomised controlled trials (RCTs) will pave the way for machine learning models entering the clinical routine. Nevertheless, not every possible choice of model architecture can be tested prospectively due to the restricted sample sizes (and therefore, number of study arms). Rather, the development of these models is generally assumed to occur retrospectively. However, precisely those retrospective studies are facing multiple obstacles, which we are going to discuss next.

### 4.2 Comparability

Concerning the comparability of the reviewed studies, we note that there are several challenges that have yet to be overcome, namely the choice of (i) prediction task, (ii) case-control onset matching, (iii) sepsis definition, (iv) implementation of a given sepsis definition, and (v) performance measures. We subsequently discuss each of these challenges.

#### 4.2.1 Prediction task

As described in Section 3.5, we found that the vast majority of the included papers follow one of two major approaches when implementing the sepsis onset prediction task: Either an offline training step was followed by a horizon evaluation, or both the training and the evaluation were conducted in an online fashion. As one of our core findings, we next highlight the strengths but also the intricacies of these two setups. Considering the most frequently-used strategy, i.e. offline training plus horizon evaluation, we found that the horizon evaluation provides valuable information about how early (in hours before sepsis onset) the machine learning model is able to recognise sepsis. However, in order to train such a classifier, the choice of a meaningful time window (and matched onset) for controls is an essential aspect of the study design (for more details, please refer to Section 4.2.2). By contrast, the online strategy does not require a matched onset for controls (see Figure 4), but it removes the convenience of easily estimating predictive performance for a given prediction horizon (i.e. in hours before sepsis onset). Nevertheless, models trained and evaluated in an online fashion may be more easily deployed in practice, as they are by construction optimised for continuously predicting sepsis while new data arrives. Meanwhile, in the offline setting, the entire classification task is retrospective because all input data are extracted right up until a previously-known sepsis onset. Whether a model trained this way would generalise to a prospective setup in terms of predicting sepsis *early* remains to be analysed in forthcoming studies. In this review, the only study featuring prospective analysis focused on (and improved) prospective targets other than sepsis onset, namely mortality, length of stay, and hospital readmission. Finally, we observed that the online setting also contains a non-obvious design choice, which is absent in the offline/horizon approach: How many hours *before* and *after* a sepsis onset should a positive prediction be considered a true positive or rather a false positive? In other words, how long before or after the onset should a model be incentivised to raise an alarm for sepsis? Reyna et al. [2019] proposed a clinical utility score that customises a clinically-motivated reward system for a given positive or negative prediction with respect to a potential sepsis onset. For example, it reflects that late true positive predictions are of little to no clinical importance, whereas late false negatives predictions can indeed be harmful. While such a hand-crafted score may account for a clinician’s diagnostic demands, the resulting score remains highly sensitive to the exact specifications for which there is currently neither an internationally-accepted standard nor a consensus. Furthermore, in its current form, the proposed clinical utility score is hard to interpret.

#### 4.2.2 Case-control onset matching

Futoma et al. [2017b] observed a drastic drop in performance upon introducing their (relative) case-control onset matching scheme as compared to an earlier version of their study, where the classification scenario compares sepsis onsets with the discharge time of controls [Futoma et al.2017a]. Such a matching can be seen as an implicit onset matching, which studies that do not account for this issue tend to default to. This suggests that comparing the data distribution of patients at the time of sepsis onset with the one of controls when being discharged could systematically underestimate the difficulty of the relevant clinical task at hand, i.e. identifying sepsis in an ICU stay. Futoma et al. [2017b] also remarked that “For non-septic patients, it is not very clinically relevant to include all data up until discharge, and compare predictions about septic encounters shortly before sepsis with predictions about non-septic encounters shortly before discharge. This task would be too easy, as the controls before discharge are likely to be clinically stable.” The choice of a matched onset time is therefore crucial and highlights the need for a more uniform reporting procedure of this aspect in the literature. Furthermore, Moor et al. [2019] proposed to match the *absolute* sepsis onset time (i.e. perform absolute onset matching) to prevent biases that could arise from systematic differences in the length of stay distribution of sepsis cases and controls (in the worst case, a model could merely re-iterate that one class has shorter stays than the other one, rather than pick up an actual signal in their time series). Lastly, Table 2 lists four studies that employed random onset matching. Given that sepsis onsets are not uniformly distributed over the length the ICU stay (for more details, please refer to Section 4.4), this strategy could result in overly distinct data distributions between sepsis cases and non-septic controls.

#### 4.2.3 Defining and implementing sepsis

A heterogeneous set of existing definitions (and modifications thereof) was implemented in the reviewed studies. The choice of sepsis definition will affect studies in terms of the prevalence of patients with sepsis and the level of difficulty of the prediction task (due to assigning earlier or later sepsis onset times). We note that it remains challenging to fully disentangle all of these factors: on the one side, a larger absolute count of septic patients is expected to be beneficial for training machine learning models (in particular deep neural networks). On the other side, including more patients could make the resulting sepsis cohort a less severe one and harder to distinguish from non-septic ICU patients. Then again, a more inclusive sepsis labelling would result in a higher prevalence (i.e. class balance), which would be beneficial for the training stability of machine learning models. To further illustrate the difficulty of defining sepsis, consider the prediction target *in-hospital mortality*. Even though in-hospital mortality rates (and therefore any subsequent prediction task) vary between cohorts and hospitals, their *definition* typically does not. Sepsis, by contrast, is inherently hard to define, which over the years has led to multiple refinements of clinical criteria (Sepsis 1-3) for trying to capture sepsis in one easy-to-follow, rule-based definition [Bone et al., 1992, Levy et al., 2003, Singer et al., 2016]. It has been previously shown that applying different sepsis definitions to the same dataset results in largely dissimilar cohorts [Johnson et al., 2018]. Furthermore, this specific study found that using Sepsis-3 is too inclusive, resulting in a large cohort showing mild symptoms. By contrast, practitioners have reported that Sepsis-3 is indeed too *restrictive* in that sepsis cannot occur without organ dysfunction any more [Johnson et al., 2018]. This suggests that even *within* a specific definition of sepsis, substantial heterogeneity and disagreement in the literature prevails. On top of that, we found that even applying the same definition on the same dataset has resulted in dissimilar cohorts. Most prominently, in Table 1, this can be confirmed for studies employing the MIMIC-III dataset. However, the determining factors cannot be easily recovered, as the code for assigning the labels is not available in 19 out of 21 (90.4%) studies employing computer-derived sepsis labels. Another factor exacerbating comparability is the heterogeneous sepsis prevalence. This is partially influenced by the training setup of a given study, because certain studies prefer balanced datasets for improving the training stability of the machine learning model [Bloch et al., 2019, Van Wyk et al., 2019, van Wyk et al., 2019], while others preserve the observed case counts to more realistically reflect how their approach would fare when being deployed in ICU. Furthermore, the exact sepsis definition used as well as the applied data pre-processing and filtering steps influence the resulting sepsis case count and therefore the prevalence [Johnson et al., 2018, Moor et al., 2019]. Figure 2 depicts a boxplot of the prevalence values of all studies. Out of the 22 studies, 10 report prevalences ≤ 10%, with the maximum reported prevalence being 63.6% [Kaji et al., 2019]. In addition, Figure 5 depicts the distribution of all sepsis encounters, while also encoding the sepsis definition (or modification thereof) that is being used.

**Figure 5:**
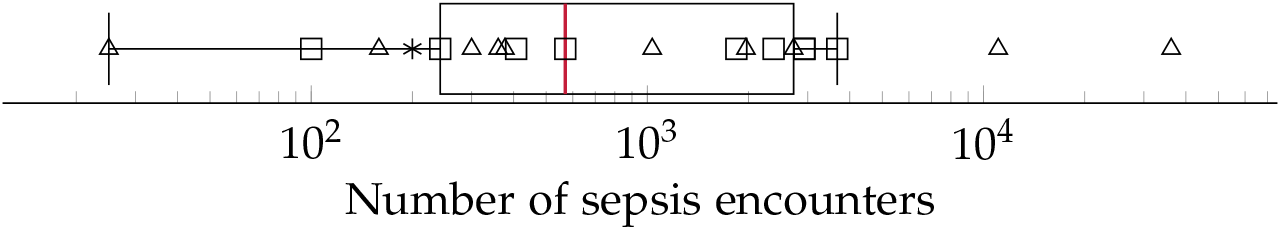
A boxplot of the number of sepsis encounters reported by all studies, with the median number of encounters being highlighted in red. Since the numbers feature different orders of magnitude, we employed logarithmic scaling. The marks indicate which definition or modification thereof was used. Sepsis-3: squares, Sepsis-2: triangles, domain expert label: asterisk.

#### 4.2.4 Performance measures

The last obstacle impeding comparability is the choice of performance measures. This is entangled with the differences in sepsis prevalence: simple metrics such as accuracy are directly impacted by class prevalence, rendering a comparison of two studies with different prevalence values moot. Some studies report the area under the receiver operating characteristic curve (AUROC, sometimes also reported as AUC). However, AUROC also depends on class prevalence and is known to be less informative if the classes are highly imbalanced [Lobo et al., 2008, Saito and Rehmsmeier, 2015]. The area under the precision-recall curve (AUPRC, sometimes also referred to as average precision) should be reported in such cases, and we observed that *n* = 6 studies already do so. AUPRC is also affected by prevalence but permits a comparison with a random baseline that merely “guesses” the label of a patient. AUROC, by contrast, can be high even for classifiers that fail to properly classify the minority class of sepsis patients. This effect is exacerbated with increasing class imbalance. Recent research suggests reporting the AUPRC of models, in particular in clinical contexts [Pinker, 2018], and we endorse this recommendation.

#### 4.2.5 Comparing studies of low comparability

Our findings indicate that quantitatively comparing studies concerned with machine learning for the prediction of sepsis in the ICU is currently a nigh-impossible task. While one would like to perform meta-analyses in these contexts to aggregate an overall trend in performance among state-of-the-art models, at the current stage of the literature this would carry little meaning. Therefore, we currently cannot ascertain the best performing approaches by merely assessing numeric results of performance measures. Rather, we had to resort to *qualitatively* assess study designs in order identify underlying biases which could lead to overly optimistic results.

### 4.3 Reproducibility

Reproducibility, i.e. the capability of obtaining similar or identical results by independently repeating the experiments described in a study, is the foundation of scientific accountability. In recent years, this foundation has been shaken by the discovery of failures to reproduce prominent studies in several disciplines [Baker, 2016]. Machine learning in general is no exception here, and despite the existence of calls to action [Crick et al., 2014], the field might face a reproducibility crisis [Hutson, 2018]. The interdisciplinary nature of digital medicine comes with additional challenges for reproducibility [Stupple et al., 2019], foremost of which is the issue of dealing with sensitive data (whereas for many theoretical machine learning papers, benchmark datasets exist), but also the issue of algorithmic details such as pre-processing. Our quality assessment highlights a lot of potential for improvement here: only two studies [Kaji et al., 2019, Moor et al., 2019], both from 2019, share their analysis code and the code for generating a “label” (to distinguish between cases or controls within the scenario of a specific paper). This amounts to less than 10% of the eligible studies. In addition, only four studies [Abromavičius et al. et al., 2019, Mao et al., 2018, Moor et al., 2019] report results on publicly-available datasets (more precisely, the datasets are available for research after accepting their terms and conditions). This finding is surprising, given the existence of high-quality, freely-accessible databases such as MIMIC-III [Johnson et al., 2016a] or eICU [Pollard et al., 2018]. An encouraging finding of our analysis is that a considerable number of studies (*n* = 6) report hyperparameter details of their models. Hyperparameter refers to any kind of parameter that is model-specific, such as the regularisation constant and the architecture of a neural network [Wu et al., 2019]. This information is crucial for everyone who attempts to reproduce computational experiments.

### 4.4 Circularity

Considering that the exact sepsis onset is usually unknown, most of the existing works have approximated a plausible sepsis onset via clinical criteria such as Sepsis-3 [Singer et al., 2016]. However, these criteria comprise a set of rules to apply to vital and laboratory measurements. Schamoni et al. [2019] pointed out that using clinical measurements for predicting a sepsis label, which was itself derived from clinical measurements, could potentially be circular (a statistical term referring to the fact that one uses the same data for the selection of a model and its subsequent analysis). This runs the risk being unable to discover unknown aspects of the data, since classifiers may just confirm existing criteria instead of helping to generate new knowledge. In the worst case, a classifier would merely reiterate the guidelines used to define sepsis *without* being able to detect patterns that permit an earlier discovery. To account for this, Schamoni and colleagues chose a questionnaire-based definition of sepsis and clinical experts manually labelled the cases and controls. While this strategy may reduce the problem of circularity, a coherent and comprehensive definition of sepsis cannot be easily guaranteed. Notably, Schamoni et al. [2019] report very high inter-rater agreement. They assign, however, only *daily* labels which is in contrast to automated Sepsis-3 labels that are typically extracted in an hourly resolution. Furthermore, it is plausible that even with clinical experts in the loop, some level of (indirect) circularity could still take place, because a clinician would also consult the patients’ vital and laboratory measurements in order to assign the sepsis tag, it would merely be less explicit. Since Schamoni et al. [2019] proposed a way to circumvent the issue of circularity, this also means that no existing work has empirically assessed the existence (or the relevance) of circularity in machine learning-based sepsis prediction. For Sepsis-3, if the standard 72 h window is used for assessing an increase in SOFA (sequential organ failure assessment score) score, i.e. starting 48 h before suspected infection time until 24 h afterwards, and if the onset happens to occur at the very end of this window, then measurements that go 72 h into the past have influenced this label. Since the SOFA score aggregates the most abnormal measurements of the preceding 24h [Vincent et al., 1996], Sepsis-3 could even “reach” 96 h into the past. Meanwhile, the distribution of onsets using Sepsis-3 tends to be highly right-skewed, as can be seen in Moor et al. [2019], where removing cases with an onset during the first 7h drastically reduced the resulting cohort size. Therefore, we conjecture that with Sepsis-3, it could be virtually impossible to strictly separate data that is used for assigning the label from data that is used for prediction, without overly reducing the resulting cohort. Finally, the relevance of an ongoing circularity may be challenged given first promising results (in terms of mortality reduction) of the first interventional studies applying machine learning for sepsis prediction prospectively [Shimabukuro et al., 2017], without explicitly accounting for circularity.

### 4.5 Limitations of this study

A limitation of this review is that our literature search was restricted to articles listed in Embase, Google Scholar, PubMed/Medline, Scopus, and Web of Science. Considering the pace at which the research in this area—in particular, in the context of machine learning—is moving forward, it is likely that the findings of the publications described in this paper will be quickly complemented by further research. The literature search also excluded grey literature (e.g. preprints, reports), the importance of which to this topic is unknown^3^, and thus might have introduced another source of search bias. The lack of studies reporting poor performance of machine learning algorithms regarding sepsis onset prediction suggests high probability of publication bias [Dickersin and Chalmers, 2011, Kirkham et al., 2010 [Joober et al., 2012]. Publication bias is likely to result in studies with more positive results being preferentially submitted and accepted for publication [Joober et al., 2012]. Finally, our review specifically focused on machine learning applications for the prediction of sepsis and severe sepsis. We therefore used a stringent search term that potentially excluded studies pursuing a classical statistical approach of early detection and sepsis prediction.

## 5 Recommendations

This section provides recommendations how to harmonise experimental designs and reporting of machine learning approaches for the early prediction of sepsis in the ICU. This harmonisation is necessary to warrant meaningful comparability and reproducibility of different machine learning models, ensure continued model development as opposed to starting from scratch, and establish benchmark models that constitute the state-of-the-art.

As outlined above, only few studies score highly with respect to reproducibility. This is concerning, as reproducibility remains one of the cornerstones of scientific progress [Stupple et al., 2019]. The lack of comparability of different studies impedes progress because a priori, it may not be clear which method is suitable for a specific scenario if different studies lack common ground (see also the aforementioned issues preventing a meta-analysis). The way out of this dilemma is to improve reproducibility of a *subset* of a given study. We suggest the following approach: (i) picking an openly-available dataset (or a subset thereof) as an additional validation site, (ii) reporting results on this dataset, and (iii) making the code for this analysis available (including models and labels). This suggestion is flexible and still enables authors to showcase their work on their respective private datasets. We suggest that code sharing—within reasonable bounds—should become the *default* for publications as modern machine learning research is increasingly driven by implementations of complex algorithms. Therefore, a prerequisite of being able to replicate the results of any study, or to use it in a comparative setting, is having access to the raw code that was used to perform the experiment. This is crucial, as any pseudocode description of an algorithm permits many different implementations with potentially different runtime behaviour and side effects. With only two studies sharing code, method development is stymied. We thus encourage authors to consider sharing their code, for example via platforms such as GitHub (https://github.com). Even sharing only parts of the code, such as the label generation process, would be helpful in many scenarios and improve comparability. The availability of numerous open source licences [Rosen, 2004] makes it possible to satisfy the constraints of most authors, including companies that want to protect their intellectual property. A recent experiment at the International Conference of Machine Learning (ICML) demonstrated that reviewers and area chairs react favourably to the inclusion of code [Chaudhuri and Salakhutdinov, 2019]. If code sharing is *not* possible, for example because of commercial interests, there is the option to share binaries, possibly using virtual machines or “containers” [Elmenreich et al., 2018]. Providing containers would satisfy all involved parties: intellectual property rights are retained but additional studies can compare their results.

### Box 1

| Recommendation | Remarks | Details |
| --- | --- | --- |
| Make code publicly available or usable | A prerequisite of being able to replicate the results of any study, or to use any model in a comparative setting, is having access to the raw code or a binary variant thereof that was used to perform the experiments. Authors are encouraged to share their code, for example via platforms such as GitHub, or their binaries using container technologies like Docker. | GitHub, Docker |
| Use external validation for the machine learning model | External validation of a classifier is crucial for assessing the model's generalisability. Several publicly-available data sources exist that can be used for this purpose. | MIMIC-II, MIMIC-III, eICU, HiRID |
| Provide exact definition of sepsis label | Implementations vary drastically in terms of prevalence and number of sepsis encounters. Thus, reporting the label generation process is essential, particularly when labels deviate from the international definitions of sepsis. For instance, when using the eICU dataset, microbiology measurements are under-reported for defining suspected infection, yet the exact modifications of sepsis implementations have not explicitly been stated [Komorowski et al., 2018]. | Provide code of how sepsis label was determined. |
| Make data available | If possible and in compliance with international data protection laws, data sources should be made accessible to bona fide researchers. There are multiple data repositories, which researchers can use to make their data accessible, while complying with data protection laws. | Harvard Dataverse, PhysioNet, Zenodo |
| Ensure comparability of models and their performances | To advance the field, it is important that researchers compare their models to existing models in order to evaluate and compare the performance across different studies. This necessitates improvements in prevalence reporting as well as the choice of different performance metrics. | Report prevalence and AUPRC in addition to other metrics. |
| Use licences for code | Licences protect the creators and the users of code. Numerous open source licences exist, making it possible to satisfy the constraints of most authors, including companies that want to protect their intellectual property. | Apache licence, BSD licences, GPL |

As for the datasets used in a study, different rules apply. While some authors suggest that peer-reviewed publications should be come with a waiver agreement for open access data [Hrynaszkiewicz and Cockerill, 2012], we are aware of the complications of sharing clinical data. We think that a reasonable middle ground can be reached by following the suggestion above, i.e. using existing benchmark datasets such as MIMIC-III [Johnson et al., 2016b] to report performance.

Moreover, we urge authors to report additional details of their experimental setup, specifically the selection of cases and controls and the label generation/calculation process. As outlined above, the case-control matching is crucial as it affects the difficulty (and thus the significance) of the prediction task. We suggest to either follow the absolute onset matching procedure [Moor et al., 2019], which is simple to implement and prevents biases caused by differences in the length of stay distribution. In any case, forthcoming work should always report their choice of case-control matching. As for the actual prediction task, given the heterogeneous prediction horizons that we observed, we suggest that authors always report performance for a horizon of 3h or 4h (in addition to any other performance metrics that are reported). This reporting should always use the area under the precision-recall curve (AUPRC) metric as it is the preferred metric for rare prevalences [Ozenne et al., 2015]. Last, we want to stress that a description of the inclusion process of patients is essential in order to ensure comparability.

## 6 Conclusions and future directions

This study performed a systematic review of publications discussing the early prediction of sepsis in the ICU by means of machine learning algorithms. Briefly, we found that the majority of the included papers investigating sepsis onset prediction in the ICU are based on data from the same centre, MIMIC-II or MIMIC-III [Johnson et al., 2016b], two versions of a high-quality, publicly-available critical care database. Despite the data agreement guidelines of MIMIC-III stating that code using MIMIC-III needs to be published (paragraph 9 of the current agreement reads “If I openly disseminate my results, I will also contribute the code used to produce those results to a repository that is open to the research community.”), only two studies [Kaji et al 2019, Moor et al., 2019] make their code available. This leaves a lot of room for improvement, which is why we recommend code (or binary) sharing (Box 1). Of 22 included studies, only *one* reflects a non-Western (i.e. neither North-American nor European) cohort, pinpointing towards a significant dataset bias in the literature (see Supplemental Table 4 for an overview of demographical information). In addition to demographic aspects such as ethnicity, differing diagnostic and therapeutic policies as well as the availability of input data for prediction are known to impact the generation of the sepsis labels. This challenge hampers additional benchmarking efforts unless more diverse cohorts are included. Moreover, since the prediction task is highly sensitive to minor changes in study specification (including, but not limited to, the sepsis definition and the case-control alignment), the majority of investigated papers do not permit a straightforward reproduction/replication and comparison of their employed cohorts and their presented prediction task. Meta-analyses are therefore impossible, as the reported metrics pertain to different, incomparable scenarios: both prevalence and case counts are highly variable, even on the same dataset, and previous work [Futoma et al., 2017b] indicated that minor changes in the experimental setup can substantially affect the difficulty of the prediction task. As a consequence, we are currently not able to identify the most predictive method for recognising sepsis early, which then ought to be further investigated in prospective trials. All in all, we found this state of the art to leave lots of room for improvement; it would be beneficial to be able to compare different models as to their generalisability, in particular when deploying machine learning algorithms in a prospective study. We see our paper as a “call to arms” for the community and hope that our recommendations are taken in the spirit of improving this task together.

## Data Availability

We make all used study data available.

https://github.com/BorgwardtLab/sepsis-prediction-review

## Author Contributions

MM, BR, and CRJ contributed substantially to the data acquisition, extraction, analysis (i.e.quality assessment), and interpretation. Furthermore, they drafted the review article. MH made substantial contributions to data interpretation (i.e.quality assessment) and participated in revising the review article critically for important intellectual content. KB made substantial contributions to the study conception and revised the review article critically for important intellectual content.

## Data Availability Statement

The data and code for this study can be found in our GitHub repository (https://githubcom/BorgwardtLab/sepsis-prediction-review).

## Footnotes

1 This includes peer-reviewed journal articles and peer-reviewed conference proceedings.

2 The dataset was not publicly available. However, with the 2019 PhysioNet Computing in Cardiology Challenge, a pre-processed dataset from Emory University Hospital has been published [Reyna et al., 2019].

3 In the machine learning community, for example, it is common practice to use preprints to disseminate knowledge about novel methods early on.

